# Simulation of Non-Pharmaceutical Interventions on COVID-19 with an Agent-based Model of Zonal Restraint

**DOI:** 10.1101/2020.06.13.20130542

**Authors:** Lindsay Álvarez, Sergio Rojas-Galeano

## Abstract

Non-Pharmaceutical Interventions (NPI) are currently the only mechanism governments can use to mitigate the impact of the COVID-19 epidemic. Similarly to the actual spread of the disease, the dynamics of the contention patterns emerging from the application of NPIs are complex and depend on interactions between people within a specific region as well as other stochastic factors associated to demographic, geographic, political and economical conditions. Agent-based models simulate microscopic rules of simultaneous spatial interactions between multiple agents within a population, in an attempt to reproduce the complex dynamics of the effect of the contention measures. In this way, it is possible to design individual behaviours along with NPI scenarios, measuring how the simulation dynamics is affected and therefore, yielding rapid insights to perform a broad assessment of the potential of composite interventions at different stages of the epidemic. In this paper we describe a model and a tool to experiment with such kind of analysis applied to a conceptual city, considering a number of widely-applied NPIs such as social distancing, case isolation, home quarantine, total lockdown, sentinel testing, mask wearing and a distinctive “zonal” enforcement measure, requiring these interventions to be applied gradually to separated enclosed districts (zones). We find that the model is able to capture emerging dynamics associated to these NPIs; besides, the zonal contention strategy yields an improvement on the mitigation impact across all scenarios of combination with individual NPIs. The model and tool are open to extensions to account for omitted or newer factors affecting the planning and design of NPIs intended to counter the late stages or forthcoming waves of the COVID-19 crisis.

## Introduction

In view of the absence of approved drugs or vaccines for COVID-19 (up to this day), Non-Pharmaceutical Interventions (NPI) are the mechanisms that public health offices around the world are using in an attempt to mitigate the impact of its epidemic ***(Lai et al., 2020***; ***Ferguson et al., 2020***). Similarly to the actual spread of the disease, the dynamics of the contention patterns obtained as a result of application of NPIs are complex and may depend not only on interactions (contacts) between people according to their individual behaviour, but many other stochastic factors associated to demographic, geographic, political and economical conditions.

One way of getting insights about the progression of the epidemics so as to allow researchers and policy-makers to take actions to control its spread, are simulation models. In this respect, there are two main approaches. The best-known approach uses macroscopic models that represent population level dynamics with causality analysis or through transition between discrete events that affect the state of the system (System Dynamics, Discrete Event Simulation). These sort of models are intended to describe the average infection dynamics and have been used in the past to study the spread of influenza pandemics ***(Ferguson et al., 2006***), and more recently to COVID-19 ***(Ferguson et al., 2020***). They are build on the basis of the classical epidemic SIR compartment model (Susceptible, Infectious, Recovered) or variations of the same, where the entire population is split in such compartments and the dynamic of the epidemics is explained by how disease evolves in each compartment rather than at the individual level. Model parameters such as compartment transition rates are calibrated with available data, whilst others such as reproduction number or NPI settings are assumed in order to simulate hypothetical scenarios, see e.g. ***Fanelli and Piazza (2020)***; ***Wilson et al. (2020)***; ***Kantner (2020)***; ***Giordano et al. (2020)***; ***Ansumali and Prakash (2020)***; ***Ferguson et al. (2020)***; ***Lai et al. (2020)***.

The other approach resorts to microscopic modelling of interactions simulated at the individual level, also known as microsimulation; here the global phenomena are observed as an emergent property of collective dynamics. Commonly referred as Individual-Based Models or IBM, this approach assumes an inherent network structure within the population, yielding contagions dependent on the interaction (or links) between individuals, whereas onset of symptoms and disease progress are modelled with probability distributions conditioned on age structures. A number of models of this kind have been also proposed recently to study the spread and contention of COVID-19 ***(Bock et al., 2020***; ***Tuomisto et al., 2020***; ***Gomez et al., 2020)***.

The latter microsimulation approach is also known as Agent-based Modelling or ABM ***(Railsback and Grimm, 2019***). Some advantages of the ABM approach to study the epidemic include their ability to model deterministic and non-deterministic decisions made by the individuals affected by interactions with other agents within a spatial vicinity, enabling a more intuitive and closely resembling the real system being simulated. In this way, uncertainty or variance arises inherently in a ABM, enabling the model to obtain non-deterministic emergencies without input parameter variation ***(Ahmed et al., 2012)***. The randomness of the spatial patterns followed by the free movement of agents, allows a broader representation of epidemiological relevant heterogeneity in the population. In addition, the ability to define particular individual traits, as well as behaviour and disease progression per individual, are useful features to study how non-deterministic effects emerge from the application of a diverse combination of NPIs.

In this paper we propose an ABM model of the COVID-19 epidemic, incorporating a number of NPIs such as social distancing, case isolation, home quarantine, total lockdown, sentinel testing, mask protection and a distinctive “zonal” restriction where these interventions can be applied to separated districts or zones of a hypothetical city. The effect of these strategies are measured in terms of morbidity, mortality, lethality -infection fatality rate (IFR) and case fatality rate (CFR)-, doubling time, reproduction number and plots of infection, recovery and deaths occurring during the simulation timeline. The main purpose of the tool is to let modellers see which NPIs or combination of NPIs can help flattening the curve of spread (mitigate) of the disease, in addition to assess the emergencies in the corresponding epidemics indicators. The details of the contemplated NPIs, model features and the developed tool are given in the following sections, along with the results of several simulation scenarios.

## Non-Pharmaceutical Interventions

NPIs are health policies intended to mitigate the effects of the spread of a new virus when vaccines or medicines are not yet available. They consist of actions recommended to the public or in some cases, enforced by the government, that affect their daily life habits. During the COVID-19 pandemic, many of these action plans have been designed and deployed at a global scale. Here we describe those that are currently incorporated into our model.

### Social Distancing

This intervention consists of maintaining a minimum physical distance between people so as to reduce the number of close interactions between infected and healthy individuals, slowing down the speed of transmission and eventual deaths. In the model, this intervention can be set on or off, and the actual amount of people willing to comply with the intervention can be defined with a parameter between 0 and 100%.

### Case Isolation

This intervention consists of confining in an enclosed facility (home) either positive tested people or sufficiently symptomatic patients that self-isolate, until they recover from the disease. In the model, this intervention can be set on or off.

### Home Quarantine

This intervention consists of confinement of relatives or housemates living in the same household of isolated cases, during their recovery. It is assumed that quarantined healthy people in the house-hold must observe strict safety protocols to avoid contagion from the isolated case. In the model, this intervention can be set on or off.

### Total Lockdown

This intervention enforces a stay-at-home policy due to partial shutdown of economy. Only some people are authorised to move around to fulfil basic needs or to support essential services. In the model, this intervention can be set on or off with a parameter of percentage of mobility permits between 5 and 95%.

### Sentinel Testing

This intervention consists of running health campaigns performing mass testing at random locations in order to detect not isolated or unconfirmed cases of either symptomatic or asymptomatic carriers. Persons whose test result positive are isolated immediately. In the model, this intervention can be set on or off, with an option to deploy the campaigns widely or restricted to zones. The stock of tests available is a parameter of the model defined as a fraction of the population between 10 and 100%. Once the campaigns run out of tests, it is possible to replenished them with a new equal supply.

### Zonal Enforcement

This intervention assumes that the hypothetical city can be divided in different districts (or zones), and then recommends to restrict movement of residents to their respective zones only. In other words, it enforces border closures between districts. The other NPIs can be applied localised to each zone. In addition, the model enables applying a novel NPI consisting of a total lockdown to the entire city with the option of gradually lifting of the curfew by individual zones. In the model, this intervention can be set on or off; besides, it features a command to unlock one zone at a time.

### Mask Protection

This intervention consists of the use of face masks as personal protective equipment to avoid spreading, or breathing in, airborne virus particles. In the model, this intervention is currently simulated as a population habit controlled by a combination of two parameters: carriers (infected people) wearing masks or not, and susceptibles (healthy people) wearing masks or not. A particular combination will have incidence in the chance of contagion when two people encounter: 3% if both are wearing masks, 8% if carrier wears masks but susceptible do not, 50% if susceptible wears masks but carrier do not, and 90% if none wear masks.

## The Agent-based Model

### Epidemics model

We build upon the compartmental *SIRE* epidemic model (*Susceptible-Infectious-Recovered-Extinct*), but we regarded the *Infectious* compartment as an extended-state ***(Lano, 2009)*** consisting of a number of conditions that we believe explain better the dynamics of the infection. These conditions are the following: *Confirmed* or not (indicating the patient will remain isolated), *Risky* or not (meaning predisposition to develop severe or critical disease), *Asymptomatic* or not (meaning patient is unaware of being a virus carrier), *Severe* or not (meaning it requires to be hospitalised to prevent death), and *Deadly* or not (indicating the patient requires Intensive Care Unit -ICU-assistance to prevent death). The schematics of the resulting extended model, that we termed *SIRE+CARDS*, is shown in Figure 1. Notice that not all of these conditions are mutually-exclusive, as they represent incremental stages of the *I* state, evolving in a manner that is explained below.

**Figure 1.**
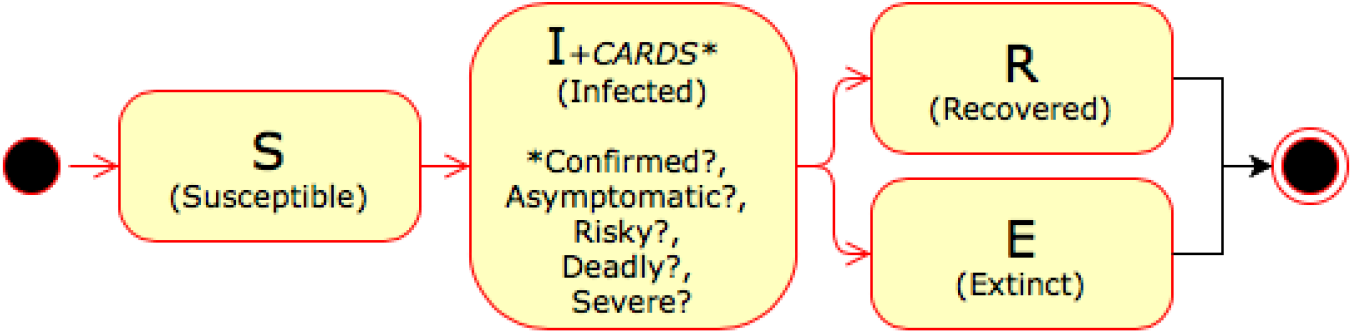
Schematic of the *SIRE+CARDS* state model.

Accordingly, we designed three types of agents: a healthy agent associated to the *S* and *R* states (the latter marked with an immunity condition), a sick agent associated to the *I* extended state tagged with the *CARDS* conditions, and a dead agent associated to the *E* state. In contrast to population-dynamic simulation, instead of using transition rates between states, our model uses transition events. A transition event is a set of decision rules evaluated on the conditions of each state. It is in this sense our proposal differs from other related ABM approaches where these conditions are regarded as additional compartments of modified SIR models, e.g. SIDHARTE ***(Giordano et al., 2020***), REINA ***(Tuomisto et al., 2020)*** or INFEKTA ***(Gomez et al., 2020)***.

Therefore, we consider the following transition events:

· *S* → *I* (infection). A healthy individual may become infected (with a probability *p-contagion*) if contact with another infectious individual occurs. When transmission happens, the new case is initially characterised as not confirmed, not tested, not severe, not deadly, not hospitalised and not ICU-admitted. Besides, a recovery (or illness) period in days is assigned as a random variate with a Gaussian distribution centered at the *avg-duration* model parameter.
· *I + Confirmed*. This condition characterises an agent as a positive case. This may happen in any of the following moments: when the patient is tested for the virus and the test results positive; when the agent has not been previously diagnosed as positive at the moment of admission to hospital, or admission to ICU bed or death; or when the patient feels sufficiently symptomatic so as to self-isolate. On the other hand, if the Case Isolation NPI (see above) is lifted after having been enforced during the course of the simulation, then the confirmed condition of agents who have not been tested positive, is reversed (i.e those who self-isolated). The distinction between confirmed and non-confirmed cases is used to identify individuals that need to stay isolated so as to prevent his capability to spread the virus. Besides, this distinction is also useful to examine the discrepancy between “official” confirmed fatality rates and the actual case fatality rate (which is computed with the totality of cases, not only the confirmed ones).
· *I + Asymptomatic*. This condition characterises an agent as not showing symptoms for the disease. The condition is activated upon acquiring the infection, with a probability defined as a model parameter (*%-asymptomatic*). In the simulation tool, this condition is indicated with the colour of the agent (yellow for asymptomatic, red for symptomatic).
· *I + Risky*. This condition characterises an agent as being in a high-risk population group. Currently the model does not consider neither risk stratification of co-morbidities nor age structure of the population. Therefore all these risk factors (obesity, diabetes, cardiovascular disease or medium to old age) are encompassed in this single condition which is activated upon acquiring the infection, with a probability defined as a model parameter (*%-high risk*).
· *I + Severity*. This condition indicates progression of the disease to a severe state, which in turn represents a higher threat of death, as it is defined in the *I* → *E* transition. This event occurs according to the following parameters (see ***Tuomisto et al. (2020)***):

| Conditions | Value |
| --- | --- |
| P(Severe \| Symptomatic, Risky) | 0.25 |
| P(Severe \| Symptomatic, no Risky) | 0.05 |
| P(Severe \| Asymptomatic) | 0.0 |

We model the day in which the severity event is triggered with a uniform distribution during the illness period: 

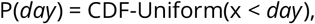

and the occurrence of severity in a given day as follows: 

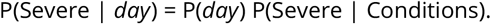

· *I + Deadliness*. This condition indicates progression of the disease to a critical state, which in turn represents an even higher threat of death, as it is defined in the *I* → *E* transition. The event occurs according to the following parameters (see ***Tuomisto et al. (2020)***):

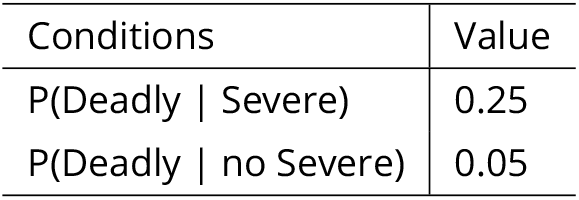

Similarly, we model the day in which the deadliness event is triggered uniformly distributed within the illness period: 

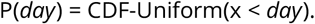

So the occurrence of deadliness in a given day is: 

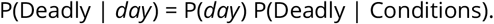

· *I + Hospitalised*. This event indicates proper care for the severity condition is taking place; hence it lowers the chance of death, as it is defined in the *I* → *E* transition. The occurrence of this event depends on the availability of *hospital-beds* (defined as a model parameter). Basically, as soon as a bed is emptied, it is assigned to an agent with severe condition in waiting (if any), until the maximum capacity is reached. When a patient is admitted, its state is switched to hospitalised, tested and confirmed (the latter two in case it was not previously diagnosed). Occupation and demand of these beds are updated on a daily basis; notice that beds are released when infectious agents recover or are translated to ICU treatment or die.
· *I + ICU-admitted*. Similar to the previous event, this one refers to proper treatment for the critical condition is being given, hence lowering the chance of death, as it is defined in the *I* → *E* transition. The assignment, discharge and availability of ICU beds is performed analogous to hospital beds, on a daily basis but considering patients with a deadly condition instead. Here, ICU beds are released when agents recover or die.
· *I* → *E* (extinction). An infected individual may become extinct with a probability that depends on his conditions according to the following parameters (see ***Tuomisto et al. (2020)***):

| Conditions | Value |
| --- | --- |
| P(Extinct \| Deadly, no ICU-admitted) | 1.0 |
| P(Extinct \| Deadly, ICU-admitted) | 0.5 |
| P(Extinct \| Severe, no Hospitalised) | 0.2 |
| P(Extinct \| Severe, Hospitalised) | 0.05 |
| P(Extinct \| Symptomatic) | 0.005 |

Since dead may occur in any day during the illness period, we modelled the chance of the day of fatality as a triangular distribution with peak in the middle of such individual’s particular recovery period (*half-recover-period*). In other words, we assume: 

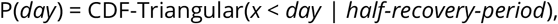

where CDF stands for cumulative distributed function. As a result, we compute the actual probability of death in a given day as follows: 

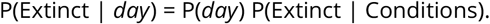

· *I* → *R* (recovery). An infected individual gets cured when he reaches the end of his recovery period. If this happens, our model assumes the individual acquires immunity to the virus and cannot be re-infected. In the simulation view, agents with this condition are coloured white.

### Agent design and behaviour

We designed agents to represent individual people residing in a hypothetical city, whose attributes include: spatial location, household location and zone or district of residence. The simulation traces an individual disease path for every agent, from susceptible to recovery or death, according to the *SIRE+CARDS* epidemic model described above. Each agent is assigned a daily routine consisting of going outdoors and returning home with a commuting distance range randomly chosen from 25, 50, 100, 200 unit steps; the actual route the agent follows varies slightly due to random fluctuations in his orientation. In addition the length of each step can be set as a global parameter between 0.1 and 1 units. Similarly, the day length can be defined with a given number of ticks in the range between 600 and 2400.

Individuals interact by random contacts they make within his/her household or during their daily routines outdoors. Virus transmission occurs when proximity within a spatial radius of 0.5 units of an infectious individual with other susceptible individuals; the chance of contagion depends on what mask protection intervention is applied, as it was explained before. Besides, our model does not consider incubation periods; infectiousness is assumed to start as soon as contagion occurs, with no difference in infectivity between symptomatic and asymptomatic carriers, or variable infectiousness between individuals. On recovery from the disease, individuals are assumed to acquire immunity to the virus so they cannot be re-infected. In this model all deaths are considered to be caused by COVID-19. No births are taken into consideration during the timeline of the simulation. A high-level depiction of the flow of events that is applied in each iteration of the simulation is shown in Figure 2. Next, we provide a brief description of each of these events.

**Figure 2.**
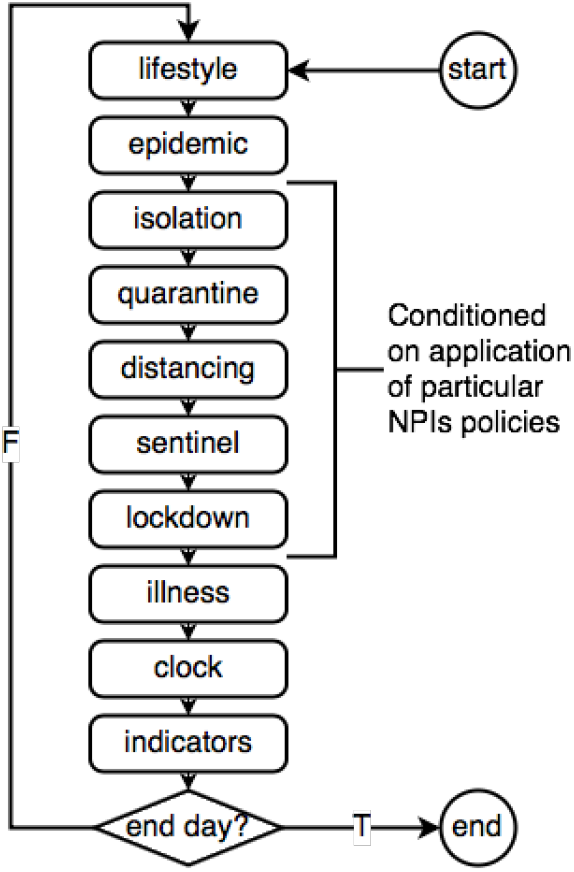
Simulation flow chart.

· **lifestyle**: simulates the daily routine of agents, which currently encompasses moving the agent ahead towards its current destination, or head it backwards home if he has reached his range limit. In any case, confirmed patients are not allowed to move around. If zonal enforcement is on, the movement is restricted only within the periphery of the zone where the agent is resident. In the simulation view, residents of different zones are identified with different shapes and colours (the latter coincides with the colour of their zone’s ground).
· **epidemic**: here the spread of the epidemic is simulated, with transmissions occurring in proportion to proximity between infectious and susceptibles and probability of contagion. If a transmission happens, the new case is initially characterised as not confirmed, not tested, not severe and not deadly. The condition of asymptomatic and/or risky of a new case is defined according to the corresponding probabilities set as model parameters. Lastly, the agent’s recovery period in days is assigned as a random variate with a Gaussian distribution centred at the *avg-duration* model parameter.
· **isolation**: when the Case Isolation NPI (CI) is enforced, those symptomatic cases that have not been tested yet are sent to their households and are tagged as confirmed cases so they have to stay isolated (their movement is restricted). If CI is lifted during the simulation, the confined cases are tagged back as not confirmed and therefore, are set free to leave its isolation.
· **quarantine**: when the Home Quarantine NPI (HQ) is enforced, housemates of confirmed cases are sent home and their movement is restricted; they will stay quarantined unless HQ is lifted during the simulation.
· **distancing**: when the Social Distancing NPI (SD) is enabled, every agent stays away of any other agent within a radius of the minimal distance model parameter. For this purpose the agent chooses the closest agent in its vicinity and heads into the opposite direction.
· **sentinel**: when the Sentinel Testing NPI (ST) is enforced, ambulances move around the entire region performing mass-testing of any agent found on their ways. Therefore each ambulance looks around its vicinity for agents that has not been tested before, perform the test on them, mark these persons as tested, and for those with positive result, sent them to isolation marked as confirmed cases. Each ambulance is supplied with a stock of tests defined as a model parameter (*%-tests*); however if they run out of tests during the simulation it is possible to replenish them with a new supply of stock. We remark that this NPI can be applied locally within each zone if the *zonal?* parameter is enabled. Lastly, notice that once a healthy agent is tested it will not be tested again in the future, unless it gets infected in which case his tested condition is reset, as it was previously explained in the epidemic event.
· **lockdown**: when the Total Lockdown NPI (TL) is enforced, all the agents are confined at home, except for a few with movement permits. The latter is intended to represent people going out for food or medicines or workers of essential jobs; they are chosen randomly every day according to a proportion of the population defined with the model parameter *%-permits*. Because our model considers zonal divisions, hence it is possible to lift the restriction in one or many zones while keeping the remainder locked down (this can be done tapping the *unlock zone* button in the simulation panel). If TL is disabled during the simulation, all the agents are unlocked so they can move freely, unless of course, they are confirmed cases. TL enforcement or lifting have effect from midnight on the next day of simulation, after having been applied.
· **illness**: in this step, progress of illness for the entire population of infectious agents is simulated according to the transition events of the *SIRE+CARDS* model described earlier. The sequence of application of these events is the following: *I* → *R, I* → *E, I + Severity, I + Deadliness, I + Hospitalised*, and lastly, *I + ICU-admitted*.
· **clock**: this event updates the day and hour counters which depends on the number of ticks (or iterations) that are needed to complete a simulated day, as defined by the corresponding model parameter.
· **indicators**: here the simulations statistics and epidemic indicators are computed, including the following:
  1. History of the number of infected people per day during the timeline of the simulation.
  2. Two estimates of the reproduction number. The first one uses a population-level approach that takes cumulative incidence data at a given day, namely the attack rate ***(Obadia et al., 2012)***; here *R*_0_ is computed as the inverse of the proportion of susceptibles after the last day of simulation of the epidemic ***(Dietz, 1993)***:

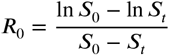

where *S*_0_ is the initial amount of susceptibles at the time of introduction of the first infected, and *S_t_* the susceptibles at time *t*. The second estimate is computed with a individual-based approach, computed directly by tracing agents, contacts and contagions individually throughout the epidemic time-line, and then taking the average: 

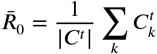

here 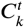 is the number of contagions made by agent *k* up to time *t*, and |*C_t_*| is the count of agents that have actually made a contagion up to that point. In this sense, instead of making assumptions about the value of reproduction number, we measure it as an emergent property of the epidemic dynamics. Both indicators, *R*_0_ and 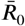 usually follow similar trends, although in some scenarios they may differ amply, as it has been previously reported ***(Breban et al., 2007)***, as it can be also verified in the corresponding plot output section of the simulation tool.
  3. The doubling time computed as ***(Bakir, 2016)***:

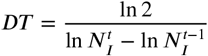

where 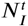 is the number of infected agents at time *t*.
  4. Additionally, the following statistics are recorded or obtained: total amount of healthy agents (susceptible or recovered), total amount of deaths, total amount of infected agents, amount of infected and confirmed agents today, new cases today, number of positive tested, and herd immunity percentage. Lethality is reported as infection fatality rate (IFR) and case fatality rate (CFR) which are computed each day as the cumulative number of deaths per cumulative number of total infections or confirmed cases, respectively.
  5. Lastly, since these indicators vary with respect to time along the duration of the simulation, we keep record of a number of plots showing the dynamics of the epidemics: SIRE plot (Susceptibles, Infectious, Recovered, Extinct), I+CARDS plot (Infectious, Confirmed, Asymptomatic, Risky, Severe, Deadly), Hospital and ICU demand plot, and R0 and Doubling Time plots.

## Simulation Tool

The simulator was developed in the NetLogo language v.6.1 (May 13, 2019). It is available as free software and can be accesed via the ModelingCommons web server in the following URL: http://modelingcommons.org/browse/one_model/6374. To experiment with the tool, choose the “Run in NetLogo Web” tab. NetLogo is a widely popular software platform for ABMs, which further to the simulation language, also integrates a graphical view area and a test-bed for experimental design ***(Wilensky and Rand, 2015)***.

The developed tool implements the NPIs, epidemic *SIRE+CARDS* model and agent behaviour rules described previously. A snapshot of the simulation view area is shown in Figure 3. In there, agents are represented with different shapes according to the zone where they reside. The colour of the agent also represents its extended state (healthy: same colour as zone ground; immune: white; sick: red, or yellow if asymptomatic; dead: black ×). Special-purpose agents designed to implement some of the NPIs such as households for home-quarantine and ambulances for sentinel-testing can also be seen.

**Figure 3.**
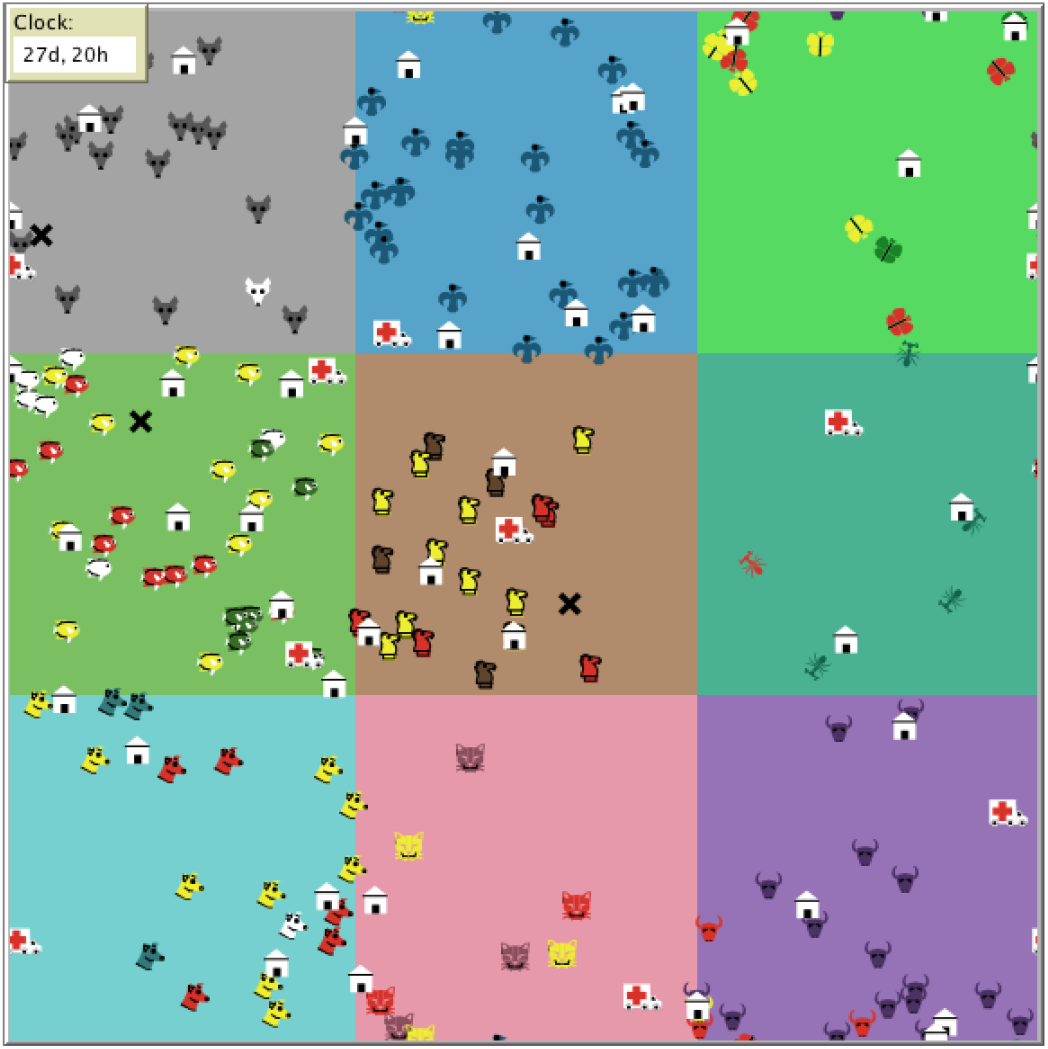
View area of the simulation tool. Agents are identified by a different shape and colour per zone; additionally, the epidemic state of agents is indicated by red (sysmptomatic), yellow (asymptomatic) or white (recovered, immune); deaths are shown by black cross marks (×). Besides, households and sentinels (ambulances) can also be seen.

The control panel is organised in sections related to general, city and COVID-19 settings, monitors of epidemic indicators, parameters, action commands to execute the simulation, and a dedicated section for NPI activation with their corresponding parameters. A snapshot of the panel is shown in Figure 4. Besides, the tool also includes another panel with plot outputs showing the dynamics of the simulation, as it was mentioned in the previous section. An example of the plots obtained in a particular execution of the model is shown in Figure 5.

**Figure 4.**
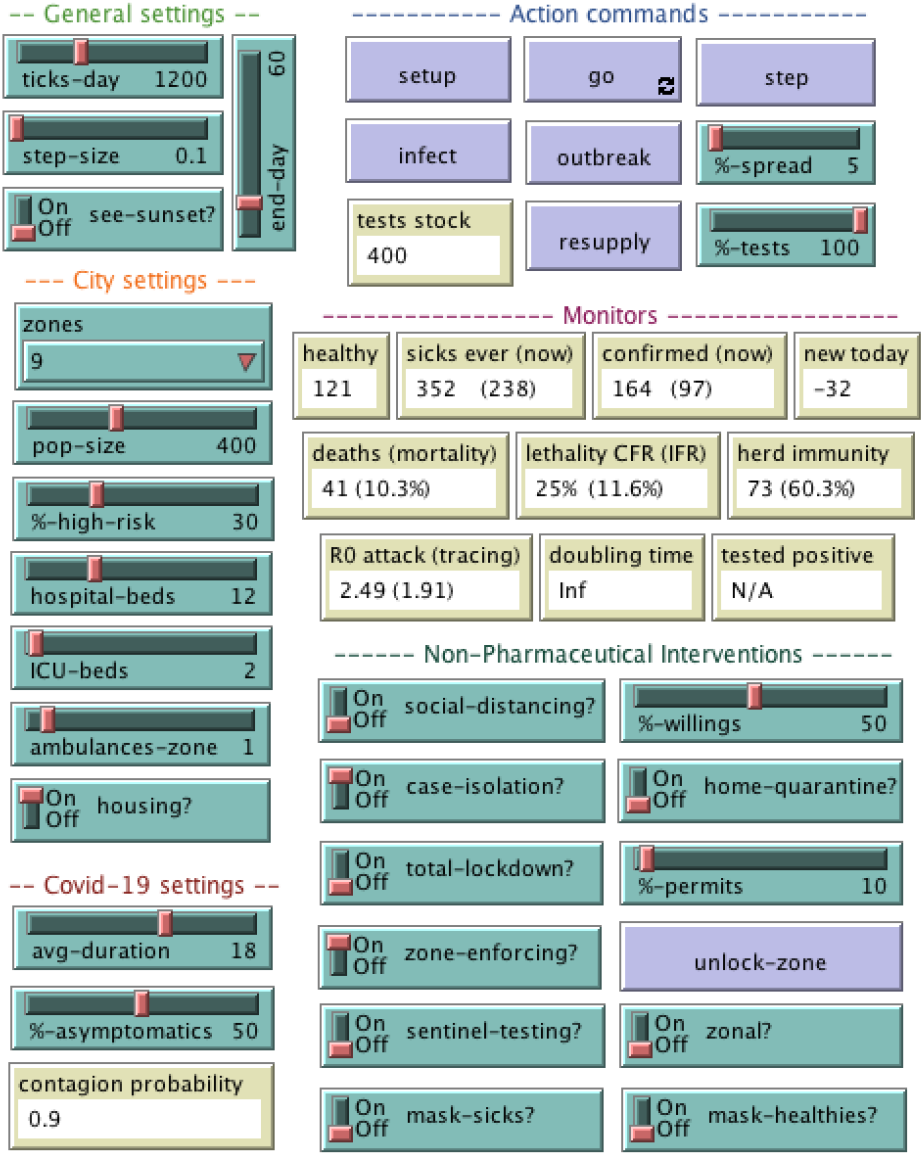
Control panel of the simulation tool.

**Figure 5.**
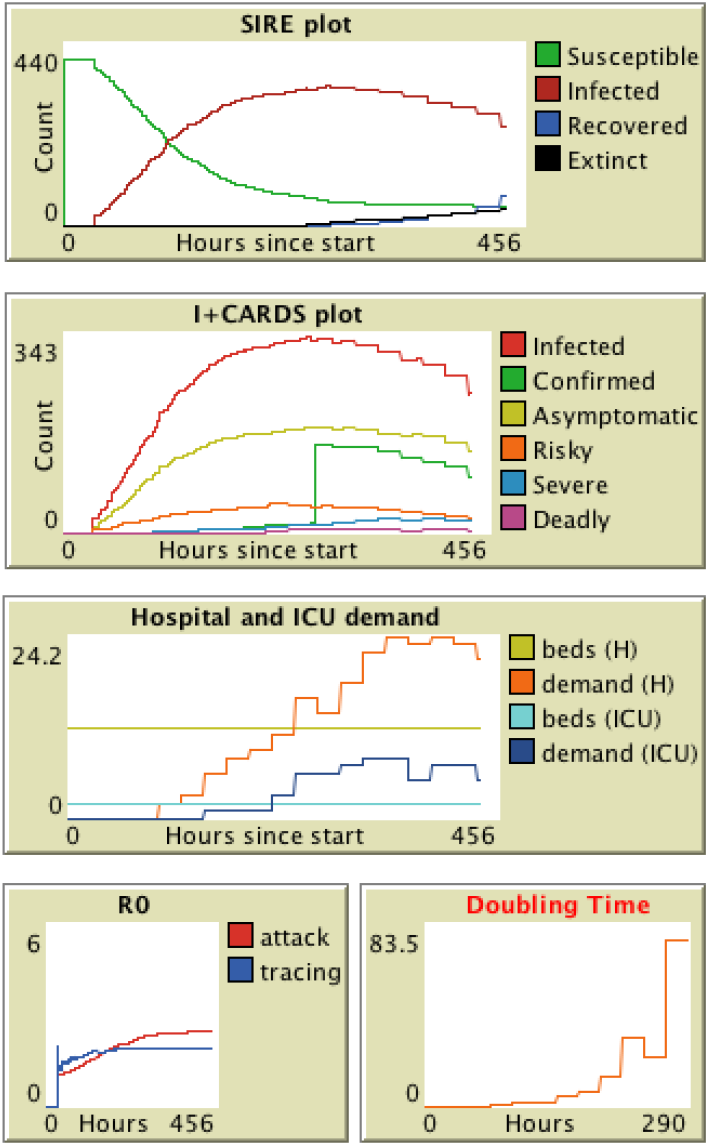
Plot area of the simulation tool.

## Hypothetical Scenarios and Results

In this section we describe a number of scenarios conceived to illustrate how to perform a rapid assessment of the effect of NPIs, or combination of NPIs, to contain the spread of the epidemics in the population of agents. We start with a baseline scenario where no measure is taken (Do Nothing). Then we simulate scenarios where the individual NPIs are applied. Finally, we simulate scenarios where these NPIs are combined with the zonal enforcement strategy in order to verify its potential impact. The description of these scenarios is given in Table 1. General, city and COVID-19 parameters used in all simulations are defined in Figure 2. For each scenario, the epidemic is assumed to begin with a “patient zero” seeded randomly in any zone of the city at 0d:12h after the start of the simulation. The application of the configured NPI policies begins at 04d:00h.

**Table 1.** Description of the simulated scenarios.

| Scenario | Description |
| --- | --- |
| DN | Baseline scenario. Patient zero is seeded at 00d:12h, epidemics unfolds with no interventions. |
| SD | Baseline plus social distancing activated with 50% of population willing to comply (%willingness). |
| CI | Baseline plus case isolation activated with no home quarantine (HQ) of relatives was applied. |
| TL | Baseline plus total lockdown with 10% of people allowed to leave households (%permits). |
| ST | Baseline plus sentinel testing with no zonal limitation. A sufficient stock of tests is guaranteed. |
| ZE | Baseline plus zone enforcement or mobility restriction within each of nine zones. |
| MP | Baseline plus mask protection (either everybody wearing masks, sick only or healthy only) |
| SD+ZE | Same as SD with zone enforcing activated. |
| CI+ZE | Same as CI with zone enforcing activated. |
| TL+ZE | Same as TL with zone enforcing activated. |
| ST+ZE | Same as ST with zone enforcing activated. |
| MP+ZE | Same as MP everybody with zone enforcing activated. |

For each scenario we performed an experiment consisting of 30 repetitions of execution of the simulation model for a timeline of 60 days. We collected the results of the count of healthy agents, deaths and immunity rate, as well as some epidemic indicators obtained at the end of the simulation time. Average results are reported in Table 3 and Table 4.

**Table 2.** Settings used in all simulated scenarios.

| Parameter | Description | Value |
| --- | --- | --- |
| pop-size | Total number of simulated people (agents) | 400 |
| zones | Number of residential zones | 9 |
| days | Period of observation days (simulation length) | 60 days |
| % high-risk | % of population with co-morbidities | 30% |
| hospital-beds | Total number of hospital beds available | 12 |
| ICU-beds | Total number of ICU beds available | 2 |
| ambulances-zone | Number of ambulances (sentinels) per zone | 1 |
| avg-duration | Average day period to recover from illness | 18 days |
| % asymptomatic | % of patients showing no or mild symptoms | 50% |

**Table 3.** Survivor, death and immune count in each scenario starting with a single infection. Statistical significant differences in counts of survivors and deaths between the single NPI scenarios compared to their corresponding combined NPI+ZE scenarios, are marked as * (p<0.01) and ** (p<0.001).

| Scenario | Survivor | Death | Immune | Immune (%) |
| --- | --- | --- | --- | --- |
| DN | 334.9 ± 8.7 | 65.1 ± 8.7 | 334.8 ± 8.8 | 1.00 ± 0.00 |
| SD | 338.8 ± 7.6 | 58.2 ± 7.3 | 329.9 ± 8.2 | 0.97 ± 0.01 |
| CI | 344.0 ± 20.0 | 55.9 ± 20.0 | 302.7 ± 102.5 | 0.90 ± 0.30 |
| TL | 344.6 ± 25.0 | 26.8 ± 14.4 | 162.1 ± 82.4 | 0.47 ± 0.24 |
| ST | 380.8 ± 13.0 | 18.8 ± 12.6 | 125.4 ± 77.7 | 0.34 ± 0.21 |
| MP sick | 329.0 ± 38.1 | 49.9 ± 20.9 | 265.2 ± 109.2 | 0.83 ± 0.33 |
| MP healthy | 334.0 ± 6.6 | 66.0 ± 6.6 | 334.0 ± 6.6 | 1.00 ± 0.00 |
| MP everyone | 361.2 ± 43.7 | 6.3 ± 9.1 | 35.0 ± 45.2 | 0.11 ± 0.15 |
| DN+ZE | 339.1 ± 49.2 | 36.5 ± 18.2 ** | 211.0 ± 104.8 | 0.65 ± 0.33 |
| SD+ZE | 356.1 ± 29.3 * | 17.9 ± 8.8 ** | 105.9 ± 48.8 | 0.31 ± 0.16 |
| CI+ZE | 375.0 ± 39.5 ** | 10.4 ± 8.2 ** | 64.0 ± 55.6 | 0.18 ± 0.16 |
| TL+ZE | 394.0 ± 3.4 ** | 6.0 ± 3.4 ** | 35.3 ± 19.7 | 0.09 ± 0.05 |
| ST+ZE | 394.8 ± 4.6 ** | 5.2 ± 4.6 ** | 34.6 ± 26.4 | 0.09 ± 0.07 |
| MP everyone +ZE | 388.8 ± 12.6 * | 5.3 ± 3.9 | 28.4 ± 20.4 | 0.07 ± 0.05 |

**Table 4.** Epidemic indicators obtained in each scenario starting with a single infection.

| Scenario | CFR (%) | IFR (%) | $R_0$<br>(attack) | $\bar{R}_0$<br>(tracing) |
| --- | --- | --- | --- | --- |
| DN | 0.69 ± 0.05 | 0.16 ± 0.02 | 21.94 ± 4.33 | 1.97 ± 0.05 |
| SD | 0.61 ± 0.05 | 0.15 ± 0.02 | 3.97 ± 0.36 | 1.87 ± 0.05 |
| CI | 0.28 ± 0.10 | 0.14 ± 0.05 | 10.15 ± 8.77 | 2.57 ± 0.88 |
| TL | 0.49 ± 0.15 | 0.11 ± 0.04 | 1.54 ± 0.34 | 2.85 ± 0.40 |
| ST | 0.10 ± 0.05 | 0.10 ± 0.05 | 1.19 ± 0.44 | 1.72 ± 0.65 |
| MP sick | 0.65 ± 0.13 | 0.21 ± 0.22 | 3.22 ± 1.25 | 1.65 ± 0.58 |
| MP healthy | 0.70 ± 0.04 | 0.17 ± 0.02 | 22.51 ± 3.12 | 1.97 ± 0.05 |
| MP everyone | 0.33 ± 0.25 | 0.07 ± 0.08 | 0.84 ± 0.58 | 1.22 ± 0.89 |
| DN+ZE | 0.56 ± 0.08 | 0.13 ± 0.03 | 3.55 ± 5.41 | 1.94 ± 0.14 |
| SD+ZE | 0.50 ± 0.10 | 0.12 ± 0.04 | 1.29 ± 0.19 | 1.93 ± 0.24 |
| CI+ZE | 0.27 ± 0.23 | 0.17 ± 0.23 | 1.03 ± 0.44 | 2.05 ± 0.94 |
| TL+ZE | 0.54 ± 0.19 | 0.17 ± 0.11 | 1.06 ± 0.03 | 2.59 ± 0.57 |
| ST+ZE | 0.12 ± 0.07 | 0.12 ± 0.07 | 1.06 ± 0.05 | 1.82 ± 0.30 |
| MP everyone +ZE | 0.59 ± 0.24 | 0.15 ± 0.15 | 0.96 ± 0.33 | 1.48 ± 0.57 |

These results hint at the effectiveness of the different NPI strategies. Across all the scenarios, the deaths count varies from 65.1 to 5.3, whereas immunity rate ranges from 100% to 7%. Regarding individual NPI scenarios, that Sentinel Testing reaches the highest count of healthy people (381± 13 in average), of which about 34% acquired immunity to the disease, while also achieving the lowest rates of mortality of approximately 4% (19±13 deaths); hence, ST seems to be the single NPI yielding the major impact in the epidemics outcome. Should authorities need to prioritise application of interventions, these assessment suggests ST should be given top priority.

Now regarding the count of healthy survivors, Mask Protection for everyone achieves the second best result (361±43) followed by Total Lockdown (345±25). This is an interesting result, taking into account that financial cost of providing population with masks can be negligible compared to the economic and social costs of maintaining lengthy lockdowns. On the other hand it is also interesting to notice that Social Distancing when 50% of population willing to comply, yields a very low impact (survivors: 339±8, deaths: 58±7, immunity: 97%), only comparable with the Do Nothing scenario (survivors: 335±9, deaths: 65.1±8.7, immunity: 100%). The latter suggests that more than half of the population should embrace distancing habits if one expect this NPI to produce any mitigation impact, maybe by reinforcing awareness through extensive coverage education campaigns. We observe that Mask Protection of healthy people only, also obtains similar figures to DN, yielding a low impact NPI. MP of sicks, in contrast, is more effective in reducing the mortality rate.

Another interesting finding related to *R*_0_, shows that Mask Protection-everyone obtained the best result (0.84), followed by Sentinel Testing (1.19), and then by Total Lockdown (1.54). On the other hand, the 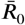 number that is computed directly by tracing the contacts of agents, yielded a best result with Mask Protection-everyone again (1.22), then Sentinel Testing (1.72), followed by Social Distancing (1.87). The latter corroborates the observation regarding the prioritisation that should be given to ST or MP interventions, plus education campaigns. Moreover, the two estimates of reproduction number can differ amply, particularly in the DN scenario (*R*_0_ = 21,94, 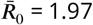), as it has been noted elsewhere ***(Breban et al., 2007)***). In view of these results, we believe the tracing approach 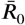 is a more realistic estimate for epidemics ABM simulations.

On the other hand, we found that the Zone Enforcement NPI exhibits interesting indicators suggesting its potential benefit for mitigation impact. Compared to the Do Nothing scenario, ZE improves death rate (from 65±9 to 37±18), CFR (from 69±5% to 56±8%) and IFR (from 16±2% to 13±3%). Since this is one of the distinctive features of our model, we thus decided to assess the effect of combining every individual NPI with the ZE restriction.

Interestingly enough, across all scenarios of NPIs combined with ZE, more effective mitigation indicators were obtained. For example, survivor and death count is better than any other scenario: TL+ZE (survivors: 394±3, death: 6±3), ST+ZE (survivors: 395±5, deaths: 5±5) and MP everyone+ZE (survivors: 389±13, deaths: 5±4). SD+ZE achieves herd immunity rate of 31%, which may be favourable for protection against a subsequent second wave of virus spread, taking into account that this scenario assumes that only 50% of population respects distancing. Regarding case fatality rate (CFR) these zone-restricted NPIs obtained ranges between 12% and 59%, whilst the infected fatality rate (IFR) obtained more optimistic values between 12% and 17%, a result that confirms the overestimation phenomenon when only official confirmed cases are accounted rather than actual contagions ***(Verity et al., 2020)***. Another observation concerning herd immunity is that following Social Distancing then comes Case Isolation (deaths: 56±20, immunity: 90±30%) and Mask Protection for sicks only (deaths: 50±21, immunity: 83±33%); then Total Lockdown (deaths: 27±14, immunity: 47±24%) and Sentinel Testing (deaths: 19±17, immunity: 34±21%).

On a different note, a noticeable pattern in these experiments is the high variability of the average results, which can be explained by two reasons. Firstly, a common theme in all these NPIs is that they perform isolation of cases as soon as they are discovered: CI by self-isolation of symptomatic cases, ST by immediate isolation of asymptomatic tested positive, and TL by mandatory confining of both symptomatic or asymptomatic (or healthy for that matter) people. Secondly, since the experiments were designed in a way that only one infected agent (patient zero) is randomly seeded at the beginning of the simulation, it is likely that in some repetitions, as soon as these NPIs take effect, patient zero can be sent by pure chance to isolation just before he or she can transmit virus to other agents; therefore, in those executions indicators may remain unaltered whilst in other repetitions the epidemics effectively unfolds fully. This kind of variability is actually an inherent property of ABM models ***(Ahmed et al., 2012)***, one that may resemble more closely the natural phenomena they simulate (take for example, the way the COVID-19 epidemics has evolved in Latin America differs amply between countries, despite sharing similar socio-economical conditions and that likewise NPIs measures have been taken by their local governments).

In order to perform an analysis ensuring the epidemics fully develops across all repetitions, we embarked in another set of experiments where rather than beginning with a single infection of a patient zero, the simulation starts with a small outbreak of 5% of the population representing “imported cases”, distributed randomly in different zones of the city. Again we run 30 repetitions per scenario for a timeline of 60 days. The average results are reported in Table 5 and Table 6.

**Table 5.** Survivor, death and immune count in each scenario starting with an outbreak of 5% population. Statistical significant differences in counts of survivors and deaths between the single NPI scenarios compared to their corresponding combined NPI+ZE scenarios, are marked as * (p<0.01) and ** (p<0.001).

| Scenario | Survivor | Death | Immune | Immune (%) |
| --- | --- | --- | --- | --- |
| DN | 334.27 ± 8.04 | 65.73 ± 8.04 | 334.20 ± 8.07 | 1.00 ± 0.00 |
| SD | 336.63 ± 7.99 | 63.30 ± 7.99 | 330.70 ± 8.00 | 0.98 ± 0.01 |
| CI | 333.57 ± 6.84 | 66.43 ± 6.84 | 332.00 ± 7.03 | 1.00 ± 0.00 |
| TL | 335.57 ± 10.23 | 64.40 ± 10.27 | 316.77 ± 11.80 | 0.94 ± 0.02 |
| ST | 353.23 ± 6.26 | 46.77 ± 6.26 | 260.27 ± 18.37 | 0.74 ± 0.05 |
| MP Sicks | 340.30 ± 8.88 | 58.60 ± 9.23 | 330.27 ± 10.06 | 0.97 ± 0.01 |
| MP Healthy | 334.63 ± 8.79 | 65.37 ± 8.79 | 334.53 ± 8.87 | 1.00 ± 0.00 |
| MP Everyone | 332.70 ± 9.45 | 33.23 ± 5.03 | 195.17 ± 29.32 | 0.59 ± 0.08 |
| DN+ZE | 336.47 ± 8.30 | 63.43 ± 8.20 | 330.97 ± 12.52 | 0.98 ± 0.04 |
| SD+ZE | 339.53 ± 11.29 | 56.83 ± 7.93 * | 322.67 ± 20.84 | 0.95 ± 0.05 |
| CI+ZE | 338.93 ± 7.55 * | 60.13 ± 8.09 * | 316.10 ± 17.31 | 0.93 ± 0.05 |
| TL+ZE | 350.80 ± 8.66 ** | 49.03 ± 8.61 ** | 276.77 ± 29.89 | 0.79 ± 0.09 |
| ST+ZE | 355.97 ± 8.99 | 44.03 ± 8.99 | 246.23 ± 21.26 | 0.69 ± 0.07 |
| MP Everyone+ZE | 345.57 ± 13.10 ** | 33.17 ± 6.35 | 194.13 ± 22.11 | 0.56 ± 0.07 |

**Table 6.**
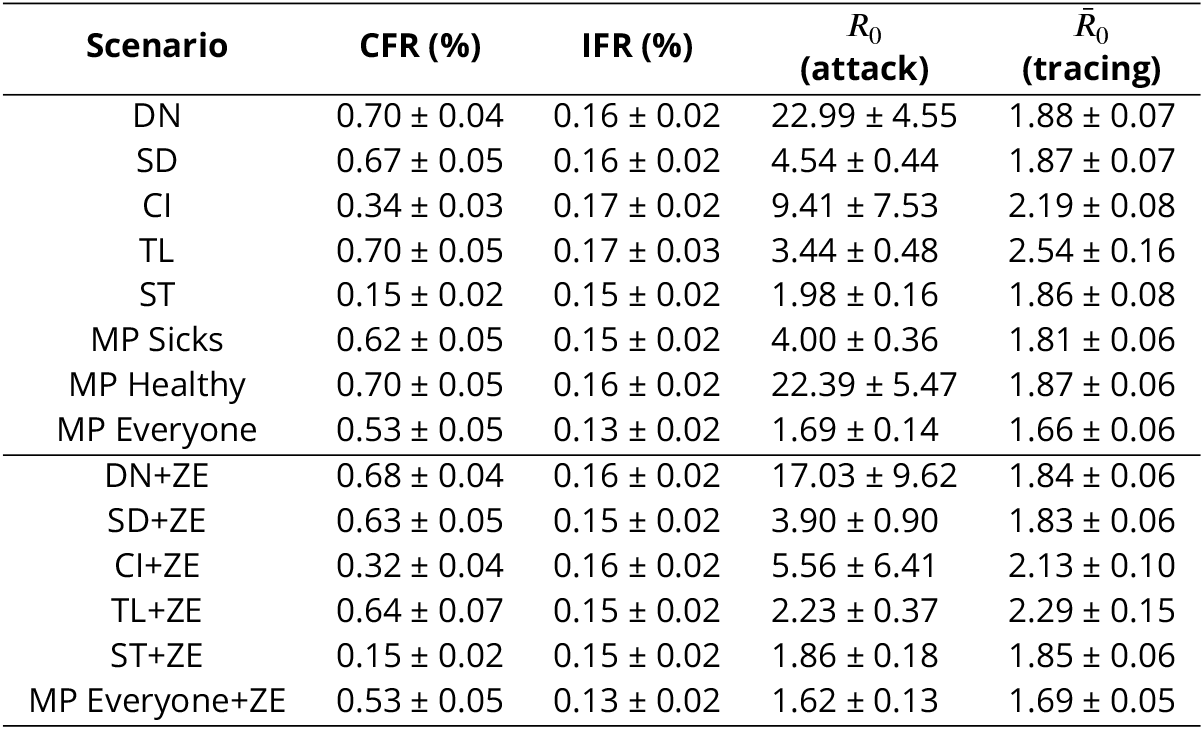
Epidemic indicators obtained in each scenario starting with an outbreak of 5% population.

| Scenario | CFR (%) | IFR (%) | $R_0$<br>(attack) | $\bar{R}_0$<br>(tracing) |
| --- | --- | --- | --- | --- |
| DN | 0.70 ± 0.04 | 0.16 ± 0.02 | 22.99 ± 4.55 | 1.88 ± 0.07 |
| SD | 0.67 ± 0.05 | 0.16 ± 0.02 | 4.54 ± 0.44 | 1.87 ± 0.07 |
| CI | 0.34 ± 0.03 | 0.17 ± 0.02 | 9.41 ± 7.53 | 2.19 ± 0.08 |
| TL | 0.70 ± 0.05 | 0.17 ± 0.03 | 3.44 ± 0.48 | 2.54 ± 0.16 |
| ST | 0.15 ± 0.02 | 0.15 ± 0.02 | 1.98 ± 0.16 | 1.86 ± 0.08 |
| MP Sicks | 0.62 ± 0.05 | 0.15 ± 0.02 | 4.00 ± 0.36 | 1.81 ± 0.06 |
| MP Healthy | 0.70 ± 0.05 | 0.16 ± 0.02 | 22.39 ± 5.47 | 1.87 ± 0.06 |
| MP Everyone | 0.53 ± 0.05 | 0.13 ± 0.02 | 1.69 ± 0.14 | 1.66 ± 0.06 |
| DN+ZE | 0.68 ± 0.04 | 0.16 ± 0.02 | 17.03 ± 9.62 | 1.84 ± 0.06 |
| SD+ZE | 0.63 ± 0.05 | 0.15 ± 0.02 | 3.90 ± 0.90 | 1.83 ± 0.06 |
| CI+ZE | 0.32 ± 0.04 | 0.16 ± 0.02 | 5.56 ± 6.41 | 2.13 ± 0.10 |
| TL+ZE | 0.64 ± 0.07 | 0.15 ± 0.02 | 2.23 ± 0.37 | 2.29 ± 0.15 |
| ST+ZE | 0.15 ± 0.02 | 0.15 ± 0.02 | 1.86 ± 0.18 | 1.85 ± 0.06 |
| MP Everyone+ZE | 0.53 ± 0.05 | 0.13 ± 0.02 | 1.62 ± 0.13 | 1.69 ± 0.05 |

The first observation in this set of outbreak experiments is that the results exhibit a smaller variability, confirming the fully unfolding intuition explained before. Moreover, compared to the patient zero experiments, the overall trend can also be identified here, except that with higher numbers. Let us examine for example, the death count here varies from 65.7 in the DN scenario to 33.2, compared to 65.1 and 5.3 in the same scenario for the patient zero experiments.

The latter increase is caused of course, by the larger amount of carriers at the beginning of the simulation (20 individuals or 5%) yielding a higher burden on the hospital facilities at the peak of the epidemics. On the contrary, immunisation also rises in the MP everyone+ZE scenario to a 56%, from a 7% in the earlier experiment. The higher levels of immunity achieved in all the scenarios are beneficial to effectively suppress the contagion or prevent it in a forthcoming second wave of the epidemics. Similar trends can be identified when comparing the other indicators with the earlier patient zero experiments.

The main finding however, in our opinion, is that simultaneous application of the Zone Enforcing intervention with the other NPIs, again yields an improvement on the mitigation impact compared to the single NPI scenarios (more survivors, fewer deaths). Significant differences between the single and their corresponding +ZE scenarios (p-value < 0.01) are highlighted in the tables. To further illustrate the effect of the application of Zone Enforcing, in the Appendix we included a comparison of the epidemic dynamics between arbitrary executions of the simulation for each pair of scenarios (single vs its corresponding combination with ZE). Particularly, the “flatenning” of the infectious curve can be seen in the +ZE plots, both as a lower height of the infectious peak and in some cases as a longer tail shifted towards a later time in hours, with respect to the start of the simulation.

## Conclusion

Given the complex nature of human behavior and virus infection, attempting to model every mechanism of the COVID-19 epidemic may prove difficult; necessary assumptions have to be made to simplify the representation of the agents and their interactions. Our aim was to focus on finding emergencies in the dynamic of this disease, considering a broad set of traits for the agents; in real life of course, there are many properties of interest that may impact the growing of the epidemics. Here we omitted detailed structures such as age, gender, profession, health status, or family, school and work clusters, transportation, job shifts or economic impacts. Clearly, those properties can be examined as extensions of the model, in the hope of capturing additional hidden patterns that may yield a better assessment of the effects NPIs can have taking additional mechanisms into consideration. Nonetheless, the results obtained indicate that the proposed model can be a useful tool in relation to a rapid assessment of the potential impact of combinations of NPIs at different stages of the contagion.

Overall, our findings suggest three relevant recommendations. In general, mass-testing (ST as we denote it in our model) is the one intervention that authorities should give priority to mitigate the impact of COVID-19, providing for this purpose with sufficient stock of tests to medical units performing random testing campaigns. Besides, Mask Protection for everyone, is even a cheaper strategy authorities should strengthen as an alternative to declaring costly lengthy total lockdowns.

In addition, the proposed zonal enforcing NPI proved to be useful to boost the mitigation impact of combination of other individual NPIs. Although this seems an appealing finding, practical application of said zonal enforcing may require logistic, residential and economical adjustments, since people usually reside and work in different districts of a city. Therefore the feasibility of limiting mobility of people within districts will depend on the urban planning and development of sufficient decentralised infrastructure, such as industrial, residential, technological, commercial and financial hubs distributed in a way that motivate people to reside and work within the same district. This may not be feasible to achieve for the current COVID-19 pandemic, particularly in cities in Latin America where proper urban zonal planning is unsatisfactory, but it is an alluring idea to start exploring as a preventive measure to counter new epidemics that may come in the forthcoming future.

Finally, interesting gaps remain to be addressed in future work. Some of the ideas we are considering are: expanding the model to include more realistic attributes regarding patients and epidemic dynamics, such as differentiated infectiousness taking into account symptomatic structures associated with age and gender; also considering incubation periods, co-morbidity risk stratification with age windows, inclusion of conglomeration centres (that is, mass transportation, schools, cinemas, hospitals), as well as the estimation of indicators of economic impacts and the study of the importance of educational aspects in the habits of collective social intelligence that may be beneficial for the mitigation power of NPIs.

## Data Availability

Experiment data is available upon request. The software is publicly available at http://modelingcommons.org/browse/one_model/6374

## Conlict of interest

The authors declare they do not have any conflict of interest.

## Appendix

Here we report plots of example executions of simulated scenarios with an outbreak infecting 5% of the population. For each scenario in Table 1, we include the individual NPIs outcome (left-hand side) and the corresponding NPIs plus zonal enforcement outcome (right-hand side).

**Figure 6.**
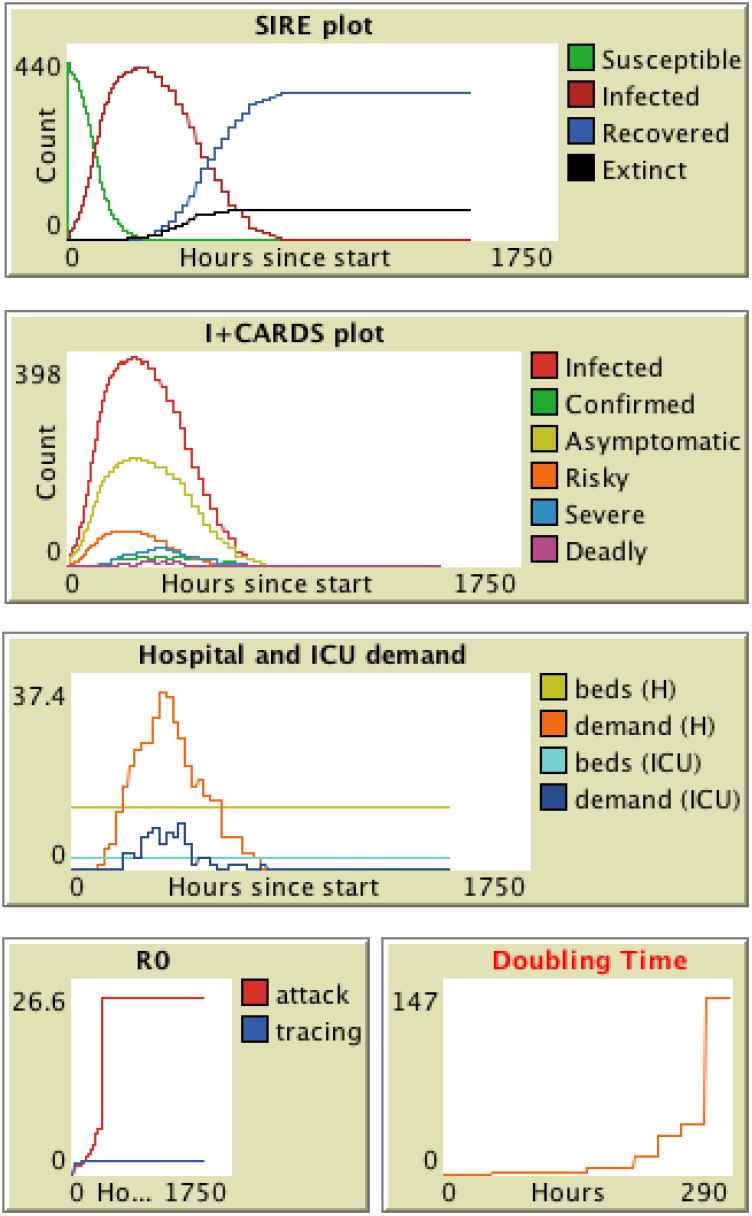
Plots of a single run of DN simulation.

**Figure 7.**
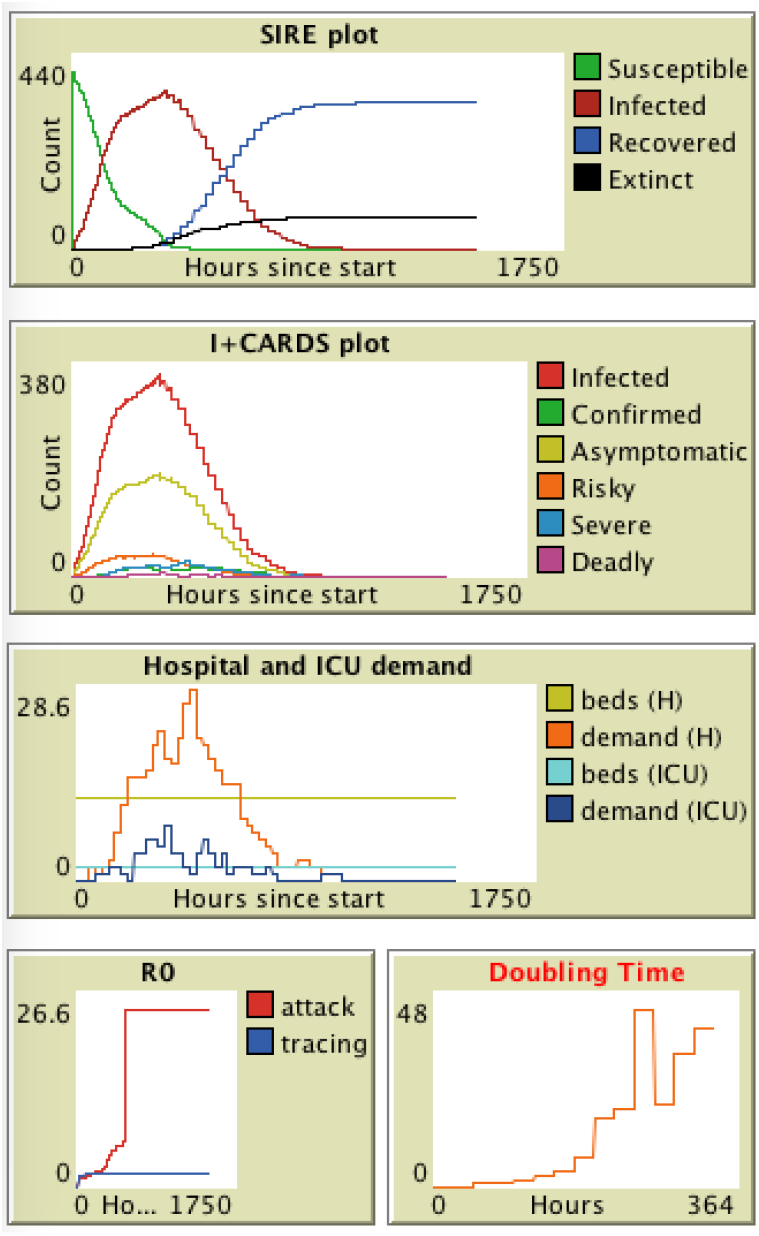
Plots of a single run of ZE simulation.

**Figure 8.**
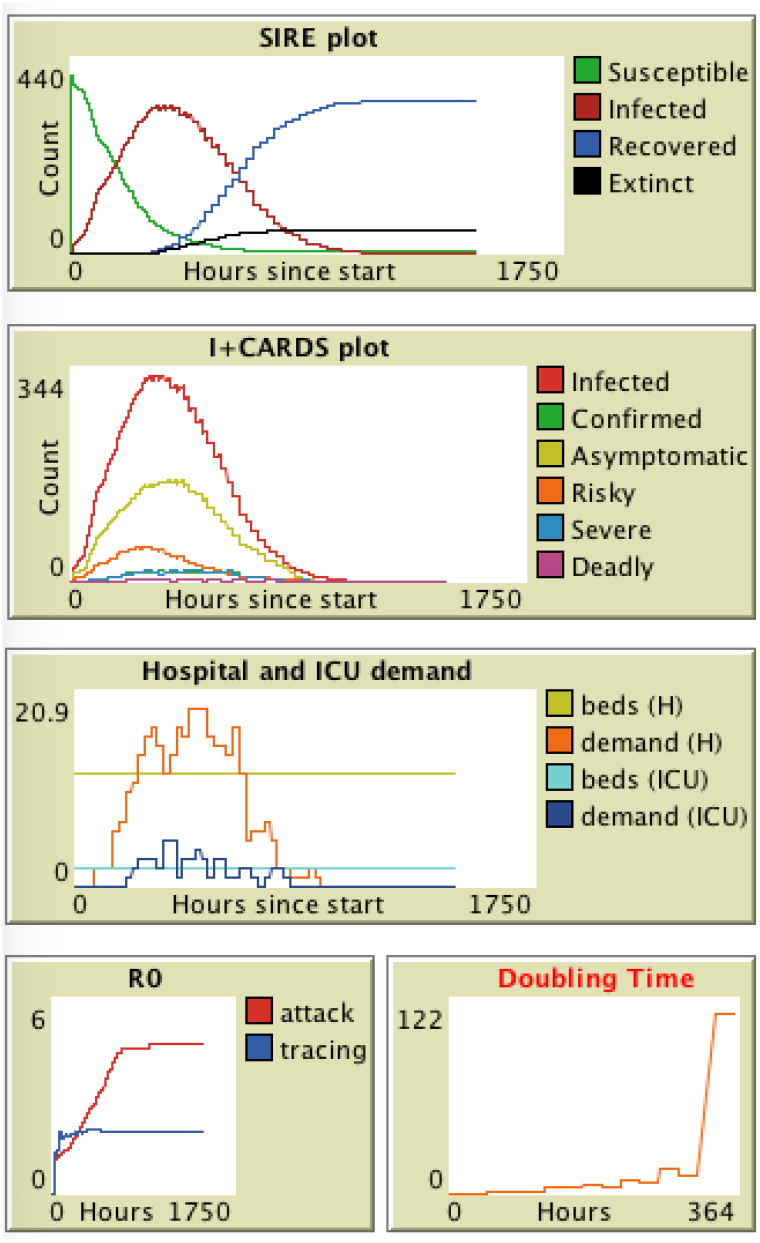
Plots of a single run of SD simulation.

**Figure 9.**
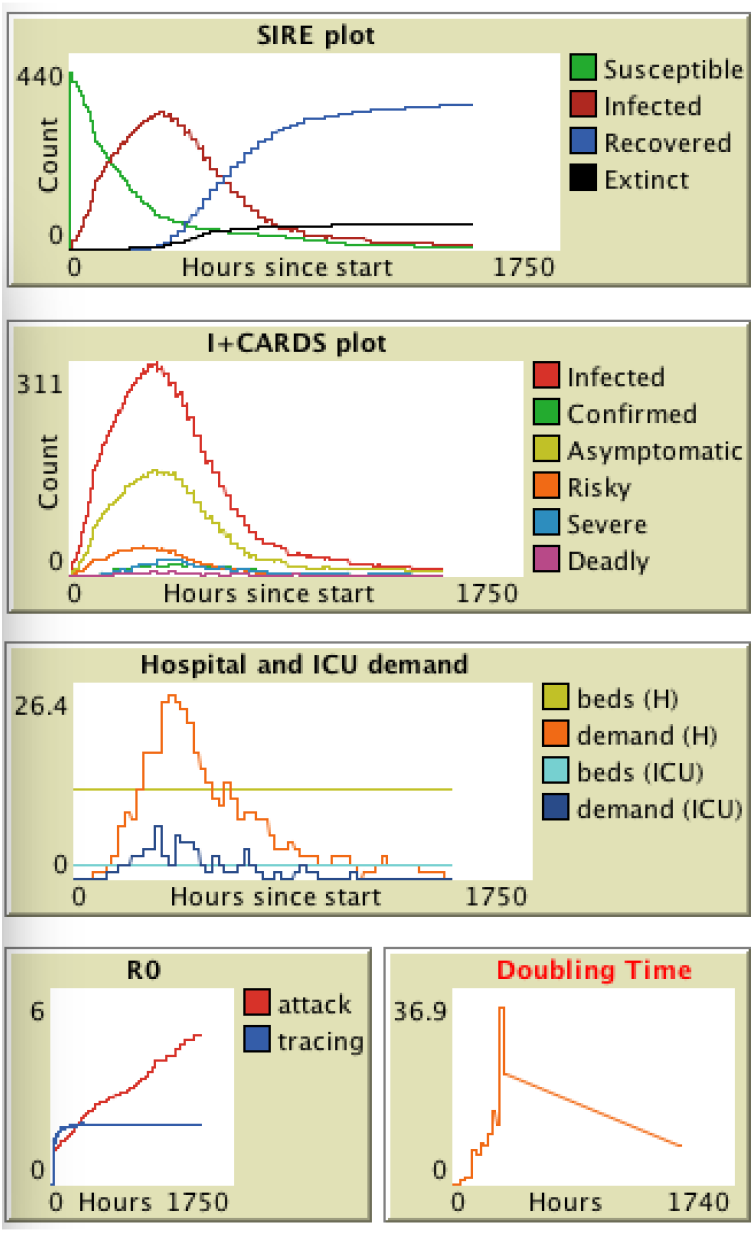
Plots of a single run of SD+ZE simulation.

**Figure 10.**
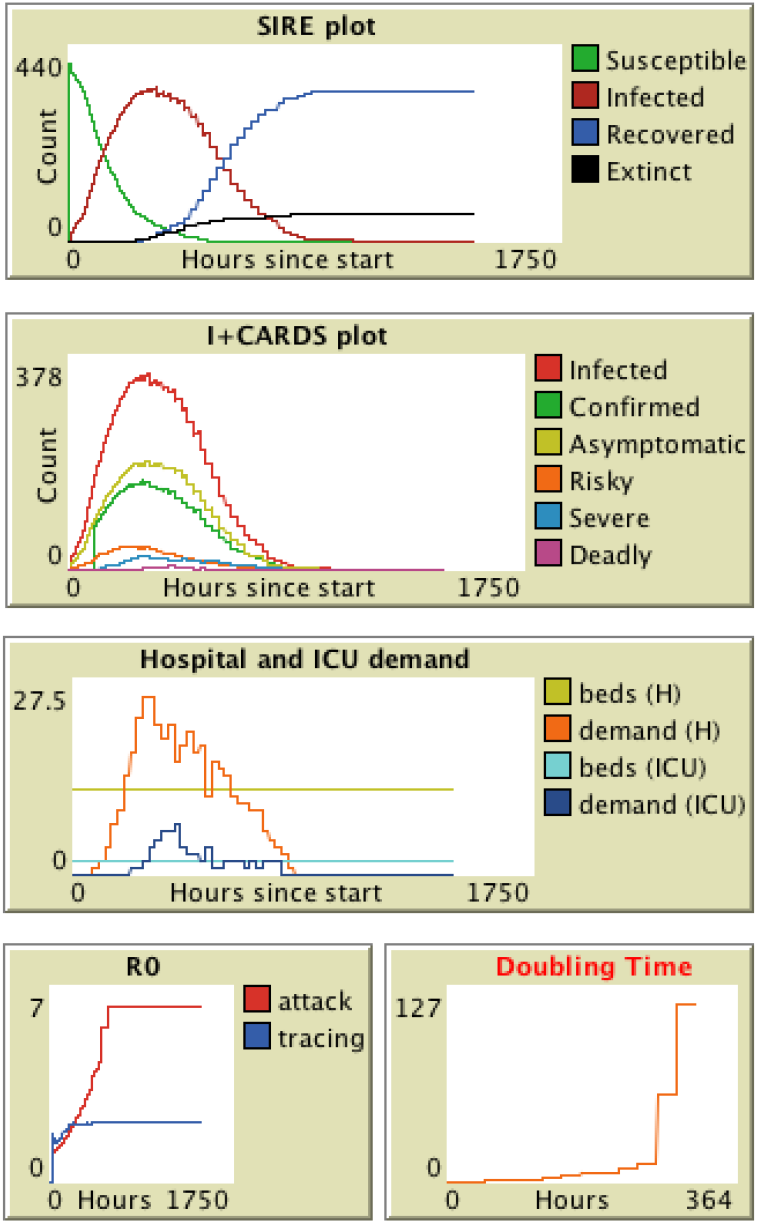
Plots of a single run of CI simulation.

**Figure 11.**
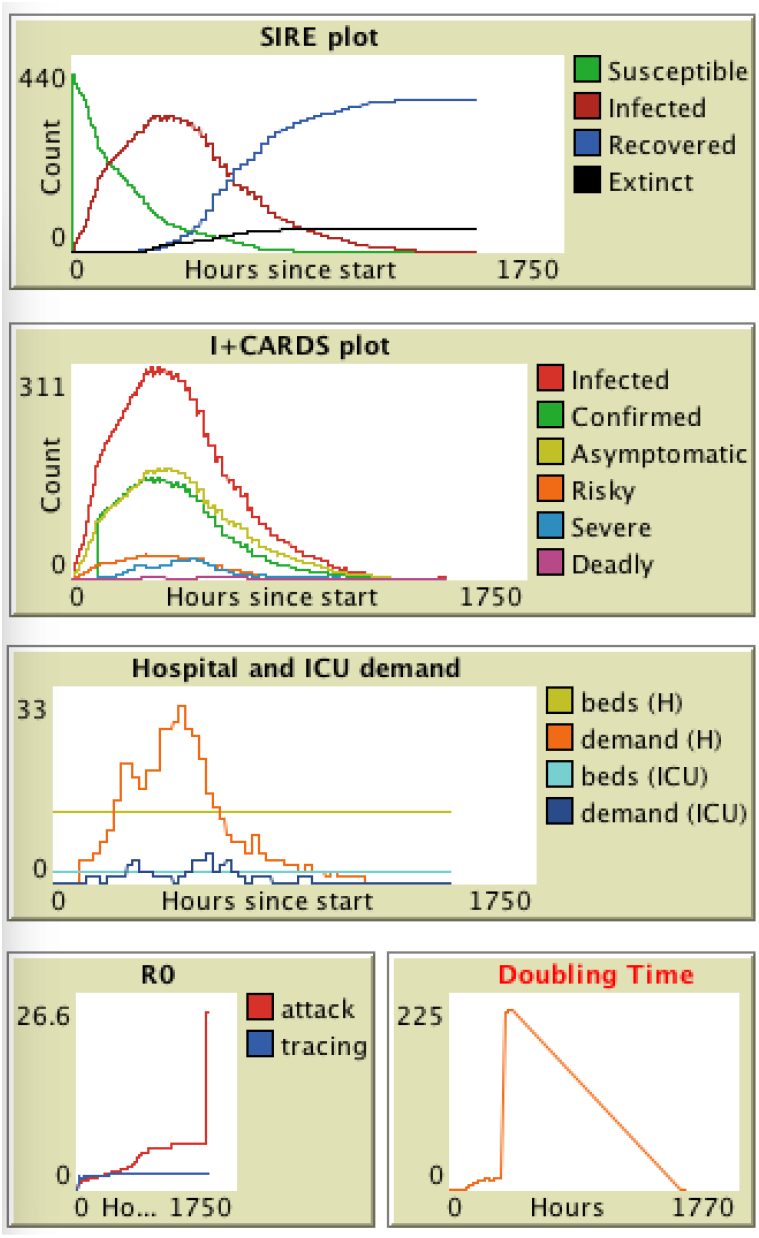
Plots of a single run of CI+ZE simulation.

**Figure 12.**
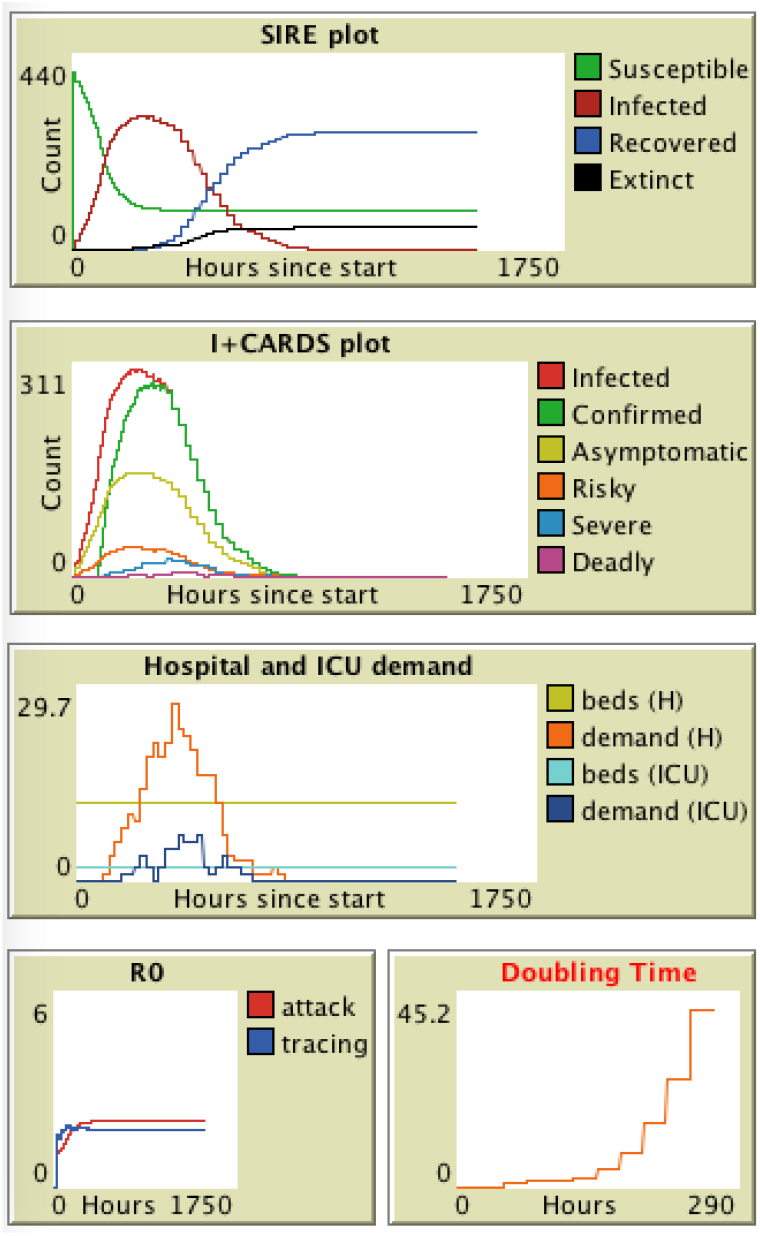
Plots of a single run of ST simulation.

**Figure 13.**
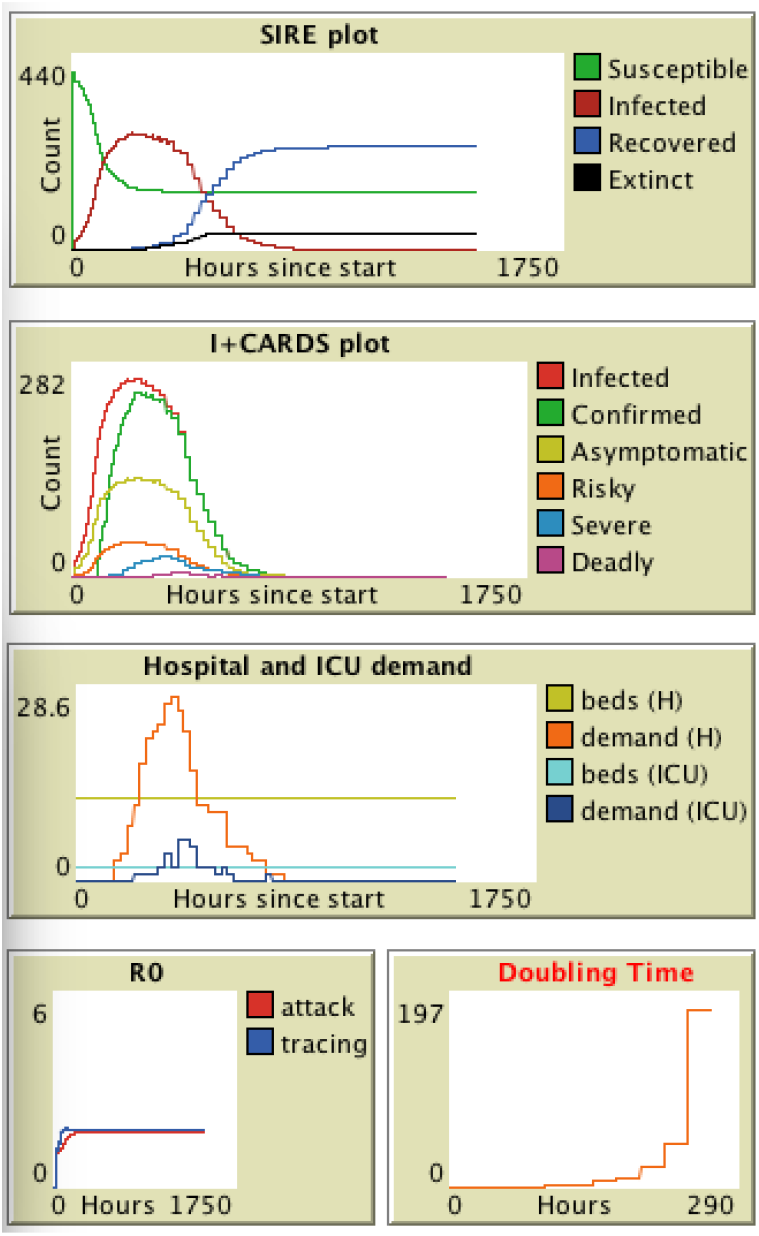
Plots of a single run of ST+ZE simulation.

**Figure 14.**
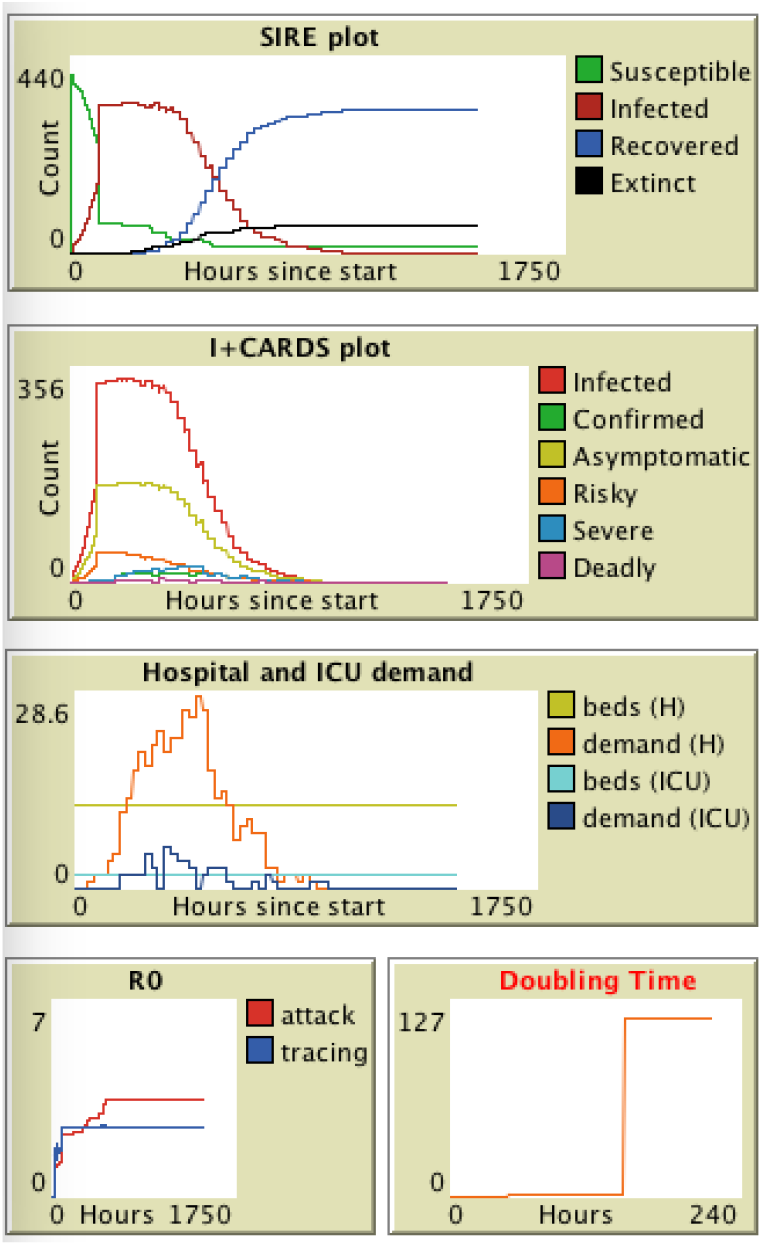
Plots of a single run of TL simulation.

**Figure 15.**
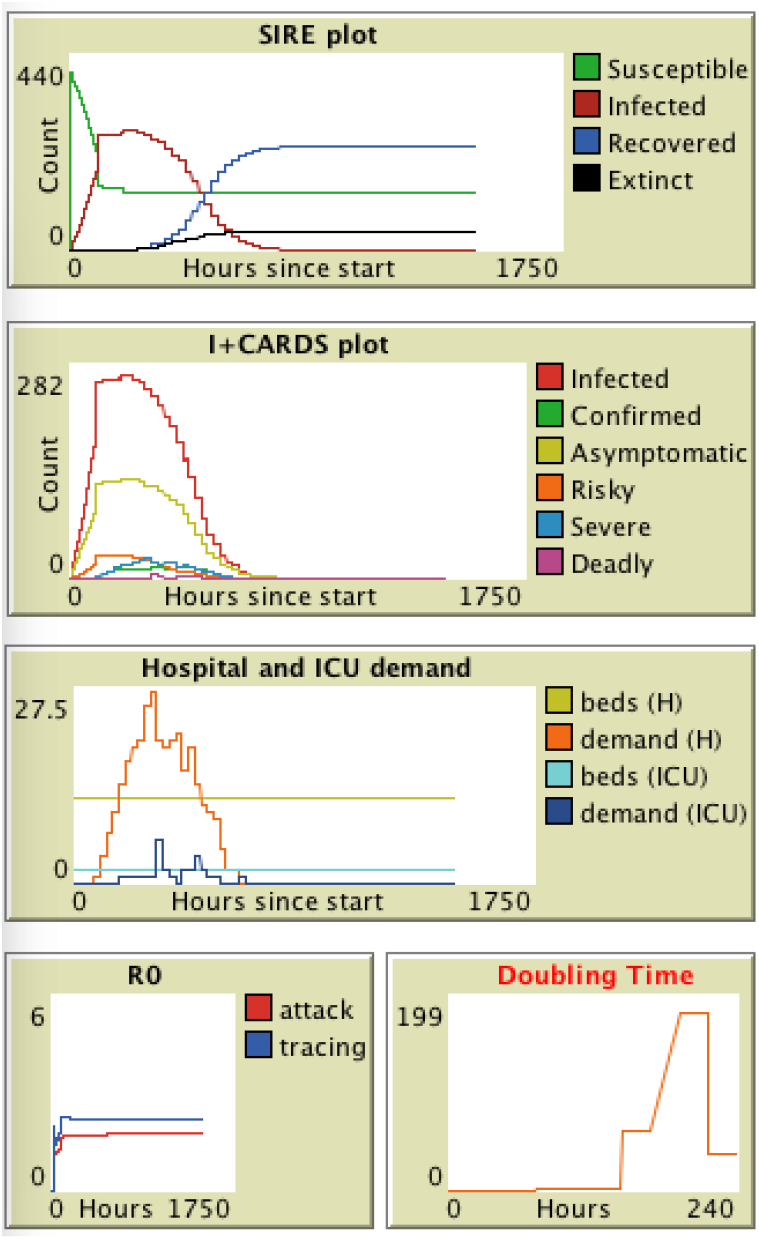
Plots of a single run of TL+ZE simulation.

**Figure 16.**
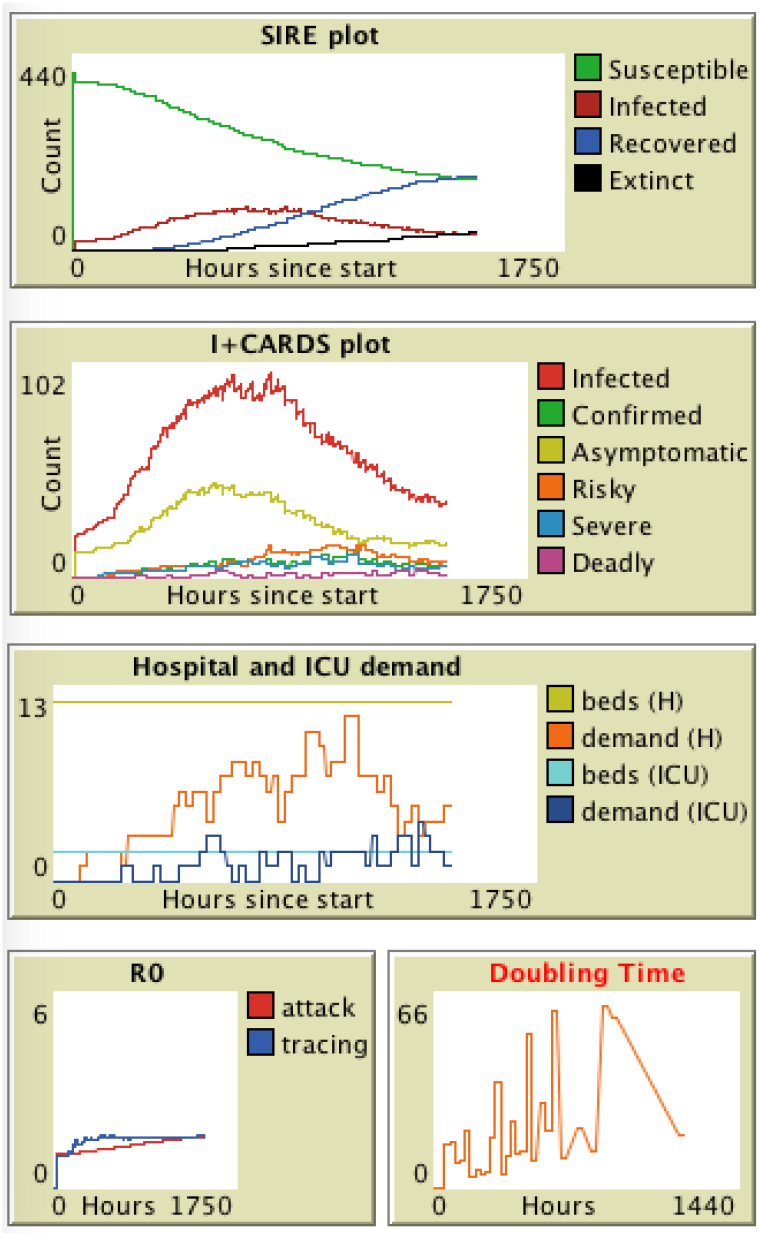
Plots of a single run of MP simulation.

**Figure 17.**
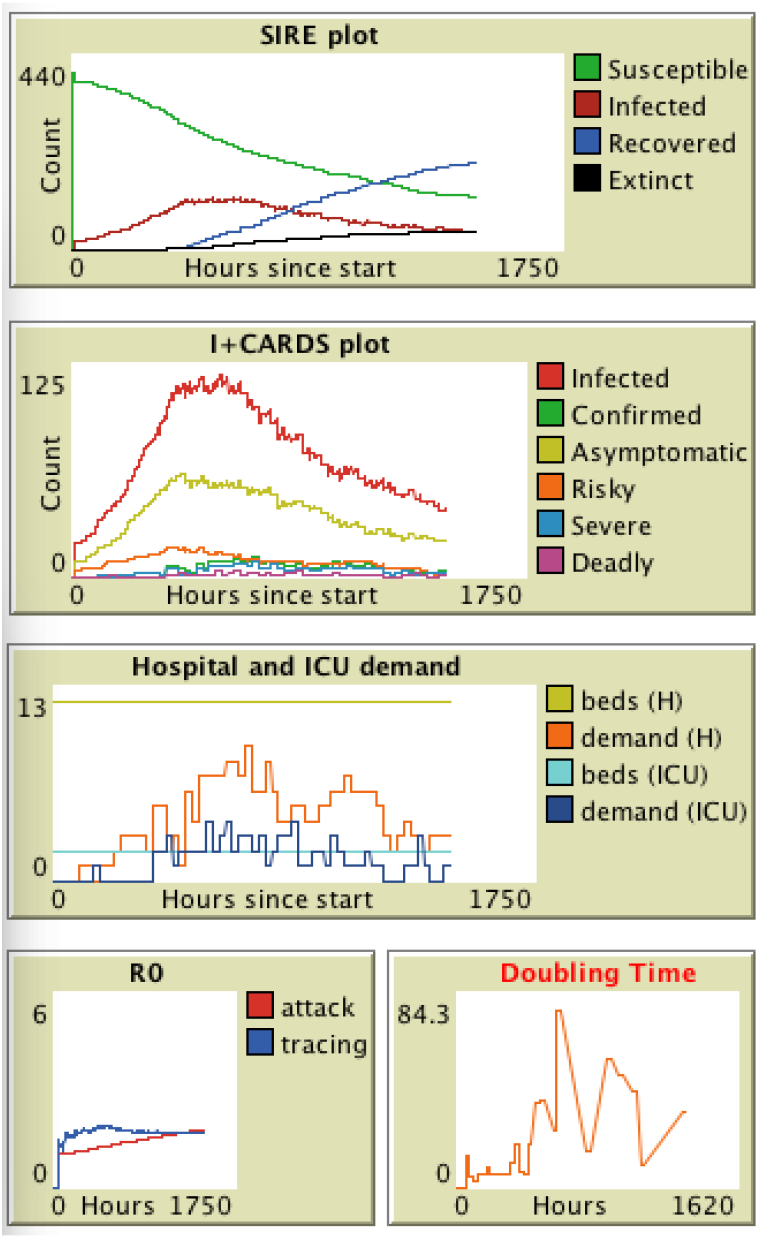
Plots of a single run of MP+ZE simulation.

